## Supplementary Materials for "Creating a Research-Ready Data Asset version of primary care data for Wales and investigating the impact of COVID-19 on utilisation of primary care services"

This supplementary material provides additional detail to support our main analyses of longitudinal coverage of linked primary care records and trends in primary care activities. It includes additional tables and figures that were not included in the main manuscript due to space constraints. These materials aim to enhance transparency and reproducibility, offering a deeper insight into our analytical approach. All results presented here align with the findings reported in the main text.

### S1. GP registration coverage

**Table S1:** Descriptive counts and column percentages of all those with records of living in Wales, every 5 years from 1990 to 2024.

|  | 1990 | 1995 | 2000 | 2005 | 2010 | 2015 | 2020 | 2024 |
| --- | --- | --- | --- | --- | --- | --- | --- | --- |
| **Total** | 2,137,980 (100%) | 2,903,630 (100%) | 2,962,900 (100%) | 3,049,370 (100%) | 3,106,980 (100%) | 3,130,210 (100%) | 3,199,040 (100%) | 3,237,480 (100%) |
| **Sex** |  |  |  |  |  |  |  |  |
| Male | 1,056,340 (49.4%) | 1,426,450 (49.1%) | 1,465,740 (49.5%) | 1,514,740 (49.7%) | 1,552,580 (50.0%) | 1,565,820 (50.0%) | 1,598,170 (50.0%) | 1,616,100 (49.9%) |
| Female | 1,081,650 (50.6%) | 1,477,180 (50.9%) | 1,497,160 (50.5%) | 1,534,640 (50.3%) | 1,554,400 (50.0%) | 1,564,390 (50.0%) | 1,600,880 (50.0%) | 1,621,380 (50.1%) |
| **Age** |  |  |  |  |  |  |  |  |
| 0-15 | 450,390 (21.1%) | 589,900 (20.3%) | 580,210 (19.6%) | 555,070 (18.2%) | 540,510 (17.4%) | 539,070 (17.2%) | 549,640 (17.2%) | 536,470 (16.6%) |
| 16-34 | 491,490 (23.0%) | 747,460 (25.7%) | 736,080 (24.8%) | 744,850 (24.4%) | 754,510 (24.3%) | 750,740 (24.0%) | 737,040 (23.0%) | 731,260 (22.6%) |
| 35-49 | 453,750 (21.2%) | 588,720 (20.3%) | 607,870 (20.5%) | 649,980 (21.3%) | 652,290 (21.0%) | 605,470 (19.3%) | 592,660 (18.5%) | 608,840 (18.8%) |
| 50-64 | 385,480 (18.0%) | 477,680 (16.5%) | 534,630 (18.0%) | 577,520 (18.9%) | 600,340 (19.3%) | 613,000 (19.6%) | 653,920 (20.4%) | 663,370 (20.5%) |
| 65-110 | 356,880 (16.7%) | 499,880 (17.2%) | 504,110 (17.0%) | 521,950 (17.1%) | 559,330 (18.0%) | 621,930 (19.9%) | 665,790 (20.8%) | 697,520 (21.5%) |
| **WIMD 2019 Quintile** |  |  |  |  |  |  |  |  |
| 1 (Most) | 455,760 (21.3%) | 621,680 (21.4%) | 603,620 (20.4%) | 606,370 (19.9%) | 612,760 (19.7%) | 623,260 (19.9%) | 644,840 (20.2%) | 654,490 (20.2%) |
| 2 | 442,430 (20.7%) | 589,710 (20.3%) | 592,870 (20.0%) | 602,200 (19.7%) | 614,220 (19.8%) | 617,100 (19.7%) | 629,930 (19.7%) | 635,650 (19.6%) |
| 3 | 432,090 (20.2%) | 589,530 (20.3%) | 603,620 (20.4%) | 621,550 (20.4%) | 633,000 (20.4%) | 635,610 (20.3%) | 649,690 (20.3%) | 658,710 (20.3%) |
| 4 | 422,990 (19.8%) | 570,040 (19.6%) | 588,200 (19.9%) | 612,470 (20.1%) | 625,590 (20.1%) | 629,440 (20.1%) | 642,140 (20.1%) | 649,430 (20.1%) |
| 5 (Least) | 384,710 (18.0%) | 532,660 (18.3%) | 574,590 (19.4%) | 606,790 (19.9%) | 621,410 (20.0%) | 624,820 (20.0%) | 632,460 (19.8%) | 639,180 (19.7%) |
| **Health Board** |  |  |  |  |  |  |  |  |
| Aneurin Bevan | 413,390 (19.3%) | 552,060 (19.0%) | 559,990 (18.9%) | 576,860 (18.9%) | 584,540 (18.8%) | 587,880 (18.8%) | 606,590 (19.0%) | 616,000 (19.0%) |
| Betsi Cadwaladr | 489,100 (22.9%) | 653,030 (22.5%) | 664,980 (22.4%) | 687,350 (22.5%) | 696,840 (22.4%) | 696,550 (22.3%) | 704,090 (22.0%) | 705,900 (21.8%) |
| Cardiff and Vale | 308,580 (14.4%) | 433,040 (14.9%) | 454,830 (15.4%) | 467,340 (15.3%) | 487,130 (15.7%) | 502,210 (16.0%) | 520,430 (16.3%) | 532,890 (16.5%) |
| Cwm Taf Morgannwg | 308,430 (14.4%) | 424,780 (14.6%) | 426,720 (14.4%) | 432,810 (14.2%) | 439,710 (14.2%) | 441,440 (14.1%) | 453,110 (14.2%) | 455,560 (14.1%) |
| Hywel Dda | 261,000 (12.2%) | 351,140 (12.1%) | 358,750 (12.1%) | 373,930 (12.3%) | 381,600 (12.3%) | 381,840 (12.2%) | 389,060 (12.2%) | 394,600 (12.2%) |
| Powys | 83,390 (3.9%) | 118,130 (4.1%) | 123,970 (4.2%) | 130,370 (4.3%) | 133,000 (4.3%) | 132,770 (4.2%) | 133,130 (4.2%) | 135,070 (4.2%) |
| Swansea Bay | 274,110 (12.8%) | 371,440 (12.8%) | 373,650 (12.6%) | 380,720 (12.5%) | 384,150 (12.4%) | 387,550 (12.4%) | 392,660 (12.3%) | 397,460 (12.3%) |

**Table S2:** Descriptive counts and column percentages of all those living in Wales with GP record linkage, every 5 years from 1990 to 2024.

|  | 1990 | 1995 | 2000 | 2005 | 2010 | 2015 | 2020 | 2024 |
| --- | --- | --- | --- | --- | --- | --- | --- | --- |
| **Total** | 752,170 (100%) | 2,285,920 (100%) | 2,522,520 (100%) | 2,687,000 (100%) | 2,750,500 (100%) | 2,762,530 (100%) | 2,796,090 (100%) | 2,792,110 (100%) |
| **Sex** |  |  |  |  |  |  |  |  |
| Male | 351,910 (46.8%) | 1,103,730 (48.3%) | 1,240,450 (49.2%) | 1,331,060 (49.5%) | 1,371,640 (49.9%) | 1,378,890 (49.9%) | 1,393,740 (49.8%) | 1,390,220 (49.8%) |
| Female | 400,260 (53.2%) | 1,182,190 (51.7%) | 1,282,070 (50.8%) | 1,355,950 (50.5%) | 1,378,860 (50.1%) | 1,383,640 (50.1%) | 1,402,360 (50.2%) | 1,401,890 (50.2%) |
| **Age** |  |  |  |  |  |  |  |  |
| 0-15 | 169,900 (22.6%) | 492,640 (21.6%) | 511,670 (20.3%) | 498,060 (18.5%) | 484,580 (17.6%) | 479,490 (17.4%) | 484,510 (17.3%) | 467,170 (16.7%) |
| 16-34 | 182,730 (24.3%) | 596,700 (26.1%) | 633,900 (25.1%) | 671,920 (25.0%) | 680,630 (24.7%) | 671,930 (24.3%) | 654,350 (23.4%) | 638,920 (22.9%) |
| 35-49 | 170,450 (22.7%) | 478,230 (20.9%) | 523,800 (20.8%) | 574,150 (21.4%) | 578,640 (21.0%) | 535,930 (19.4%) | 519,640 (18.6%) | 526,640 (18.9%) |
| 50-64 | 133,580 (17.8%) | 377,490 (16.5%) | 452,030 (17.9%) | 501,850 (18.7%) | 524,940 (19.1%) | 535,530 (19.4%) | 565,380 (20.2%) | 567,580 (20.3%) |
| 65-110 | 95,520 (12.7%) | 340,870 (14.9%) | 401,110 (15.9%) | 441,020 (16.4%) | 481,700 (17.5%) | 539,640 (19.5%) | 572,220 (20.5%) | 591,780 (21.2%) |
| **WIMD Quintile** |  |  |  |  |  |  |  |  |
| 1 (Most) | 166,980 (22.2%) | 496,180 (21.7%) | 523,740 (20.8%) | 547,970 (20.4%) | 558,330 (20.3%) | 569,180 (20.6%) | 583,140 (20.9%) | 583,750 (20.9%) |
| 2 | 163,980 (21.8%) | 474,920 (20.8%) | 515,670 (20.4%) | 545,220 (20.3%) | 558,950 (20.3%) | 556,510 (20.1%) | 562,140 (20.1%) | 558,000 (20.0%) |
| 3 | 146,420 (19.5%) | 456,860 (20.0%) | 506,320 (20.1%) | 539,680 (20.1%) | 553,400 (20.1%) | 554,200 (20.1%) | 561,460 (20.1%) | 561,730 (20.1%) |
| 4 | 141,800 (18.9%) | 428,990 (18.8%) | 476,750 (18.9%) | 510,090 (19.0%) | 522,740 (19.0%) | 524,670 (19.0%) | 530,140 (19.0%) | 529,340 (19.0%) |
| 5 (Least) | 132,990 (17.7%) | 428,970 (18.8%) | 500,040 (19.8%) | 544,050 (20.2%) | 557,080 (20.3%) | 557,980 (20.2%) | 559,220 (20.0%) | 559,280 (20.0%) |
| **Health Board** |  |  |  |  |  |  |  |  |
| Aneurin Bevan | 134,670 (17.9%) | 422,120 (18.5%) | 465,780 (18.5%) | 499,050 (18.6%) | 506,940 (18.4%) | 508,910 (18.4%) | 518,390 (18.5%) | 518,780 (18.6%) |
| Betsi Cadwaladr | 160,430 (21.3%) | 478,680 (20.9%) | 535,260 (21.2%) | 579,740 (21.6%) | 589,090 (21.4%) | 588,890 (21.3%) | 593,110 (21.2%) | 585,020 (21.0%) |
| Cardiff and Vale | 132,040 (17.6%) | 363,190 (15.9%) | 410,220 (16.3%) | 434,520 (16.2%) | 453,920 (16.5%) | 465,640 (16.9%) | 473,450 (16.9%) | 479,870 (17.2%) |
| Cwm Taf Morgannwg | 175,380 (23.3%) | 374,480 (16.4%) | 403,650 (16.0%) | 424,090 (15.8%) | 435,220 (15.8%) | 439,290 (15.9%) | 452,050 (16.2%) | 454,570 (16.3%) |
| Hywel Dda | 71,820 (9.5%) | 268,820 (11.8%) | 297,980 (11.8%) | 316,790 (11.8%) | 328,120 (11.9%) | 324,830 (11.8%) | 323,440 (11.6%) | 324,480 (11.6%) |
| Powys | 8,720 (1.2%) | 53,670 (2.3%) | 59,480 (2.4%) | 63,400 (2.4%) | 64,120 (2.3%) | 62,120 (2.2%) | 60,060 (2.1%) | 58,910 (2.1%) |
| Swansea Bay | 69,110 (9.2%) | 324,950 (14.2%) | 350,140 (13.9%) | 369,430 (13.7%) | 373,090 (13.6%) | 372,870 (13.5%) | 375,600 (13.4%) | 370,490 (13.3%) |

**Table S3:** Percentages of all those living in Wales with GP record linkage, every 5 years from 1990 to 2024.

|  | 1990 | 1995 | 2000 | 2005 | 2010 | 2015 | 2020 | 2024 |
| --- | --- | --- | --- | --- | --- | --- | --- | --- |
| **Total** | 2,137,980 (35.2%) | 2,903,630 (78.7%) | 2,962,900 (85.1%) | 3,049,370 (88.1%) | 3,106,980 (88.5%) | 3,130,210 (88.3%) | 3,199,040 (87.4%) | 3,237,480 (86.2%) |
| **Sex** |  |  |  |  |  |  |  |  |
| Male | 1,056,340 (33.3%) | 1,426,450 (77.4%) | 1,465,740 (84.6%) | 1,514,740 (87.9%) | 1,552,580 (88.3%) | 1,565,820 (88.1%) | 1,598,170 (87.2%) | 1,616,100 (86.0%) |
| Female | 1,081,650 (37.0%) | 1,477,180 (80.0%) | 1,497,160 (85.6%) | 1,534,640 (88.4%) | 1,554,400 (88.7%) | 1,564,390 (88.4%) | 1,600,880 (87.6%) | 1,621,380 (86.5%) |
| **Age** |  |  |  |  |  |  |  |  |
| 0-15 | 450,390 (37.7%) | 589,900 (83.5%) | 580,210 (88.2%) | 555,070 (89.7%) | 540,510 (89.7%) | 539,070 (88.9%) | 549,640 (88.2%) | 536,470 (87.1%) |
| 16-34 | 491,490 (37.2%) | 747,460 (79.8%) | 736,080 (86.1%) | 744,850 (90.2%) | 754,510 (90.2%) | 750,740 (89.5%) | 737,040 (88.8%) | 731,260 (87.4%) |
| 35-49 | 453,750 (37.6%) | 588,720 (81.2%) | 607,870 (86.2%) | 649,980 (88.3%) | 652,290 (88.7%) | 605,470 (88.5%) | 592,660 (87.7%) | 608,840 (86.5%) |
| 50-64 | 385,480 (34.7%) | 477,680 (79.0%) | 534,630 (84.6%) | 577,520 (86.9%) | 600,340 (87.4%) | 613,000 (87.4%) | 653,920 (86.5%) | 663,370 (85.6%) |
| 65-110 | 356,880 (26.8%) | 499,880 (68.2%) | 504,110 (79.6%) | 521,950 (84.5%) | 559,330 (86.1%) | 621,930 (86.8%) | 665,790 (85.9%) | 697,520 (84.8%) |
| **WIMD 2019 Quintile** |  |  |  |  |  |  |  |  |
| 1 (Least) | 455,760 (36.6%) | 621,680 (79.8%) | 603,620 (86.8%) | 606,370 (90.4%) | 612,760 (91.1%) | 623,260 (91.3%) | 644,840 (90.4%) | 654,490 (89.2%) |
| 2 | 442,430 (37.1%) | 589,710 (80.5%) | 592,870 (87.0%) | 602,200 (90.5%) | 614,220 (91.0%) | 617,100 (90.2%) | 629,930 (89.2%) | 635,650 (87.8%) |
| 3 | 432,090 (33.9%) | 589,530 (77.5%) | 603,620 (83.9%) | 621,550 (86.8%) | 633,000 (87.4%) | 635,610 (87.2%) | 649,690 (86.4%) | 658,710 (85.3%) |
| 4 | 422,990 (33.5%) | 570,040 (75.3%) | 588,200 (81.1%) | 612,470 (83.3%) | 625,590 (83.6%) | 629,440 (83.4%) | 642,140 (82.6%) | 649,430 (81.5%) |
| 5 (Most) | 384,710 (34.6%) | 532,660 (80.5%) | 574,590 (87.0%) | 606,790 (89.7%) | 621,410 (89.6%) | 624,820 (89.3%) | 632,460 (88.4%) | 639,180 (87.5%) |
| **Health Board** |  |  |  |  |  |  |  |  |
| Aneurin Bevan | 413,390 (32.6%) | 552,060 (76.5%) | 559,990 (83.2%) | 576,860 (86.5%) | 584,540 (86.7%) | 587,880 (86.6%) | 606,590 (85.5%) | 616,000 (84.2%) |
| Betsi Cadwaladr | 489,100 (32.8%) | 653,030 (73.3%) | 664,980 (80.5%) | 687,350 (84.3%) | 696,840 (84.5%) | 696,550 (84.5%) | 704,090 (84.2%) | 705,900 (82.9%) |
| Cardiff and Vale | 308,580 (42.8%) | 433,040 (83.9%) | 454,830 (90.2%) | 467,340 (93.0%) | 487,130 (93.2%) | 502,210 (92.7%) | 520,430 (91.0%) | 532,890 (90.1%) |
| Cwm Taf Morgannwg | 308,430 (56.9%) | 424,780 (88.2%) | 426,720 (94.6%) | 432,810 (98.0%) | 439,710 (99.0%) | 441,440 (99.5%) | 453,110 (99.8%) | 455,560 (99.8%) |
| Hywel Dda | 261,000 (27.5%) | 351,140 (76.6%) | 358,750 (83.1%) | 373,930 (84.7%) | 381,600 (86.0%) | 381,840 (85.1%) | 389,060 (83.1%) | 394,600 (82.2%) |
| Powys | 83,390 (10.5%) | 118,130 (45.4%) | 123,970 (48.0%) | 130,370 (48.6%) | 133,000 (48.2%) | 132,770 (46.8%) | 133,130 (45.1%) | 135,070 (43.6%) |
| Swansea Bay | 274,110 (25.2%) | 371,440 (87.5%) | 373,650 (93.7%) | 380,720 (97.0%) | 384,150 (97.1%) | 387,550 (96.2%) | 392,660 (95.7%) | 397,460 (93.2%) |

**Figure S1:** Mid-year population of Wales by SAIL-GP registration status from 1990 to 2024, stratified by sex.

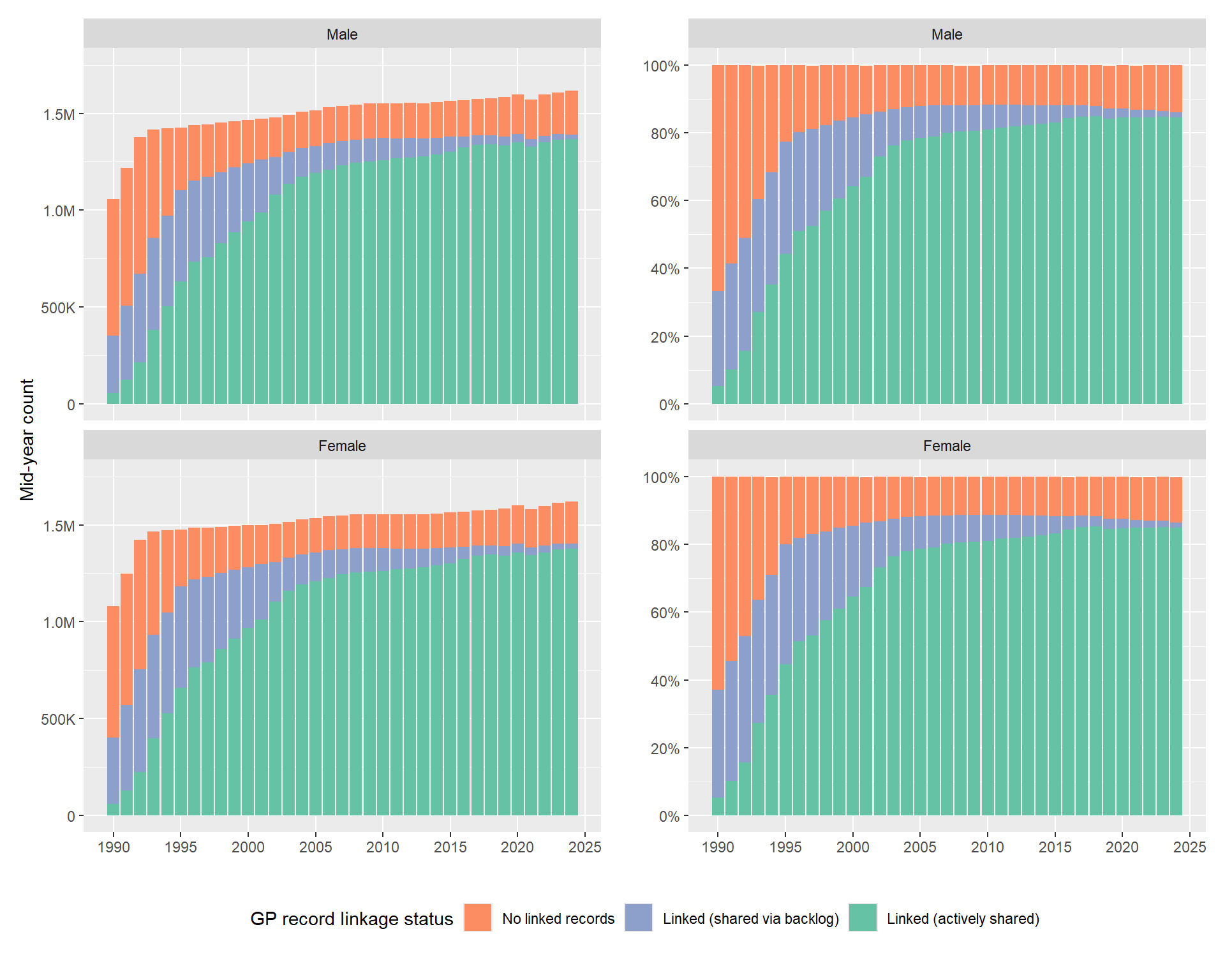

**Figure S2:** Mid-year population of Wales by SAIL-GP registration status from 1990 to 2024, stratified by age.

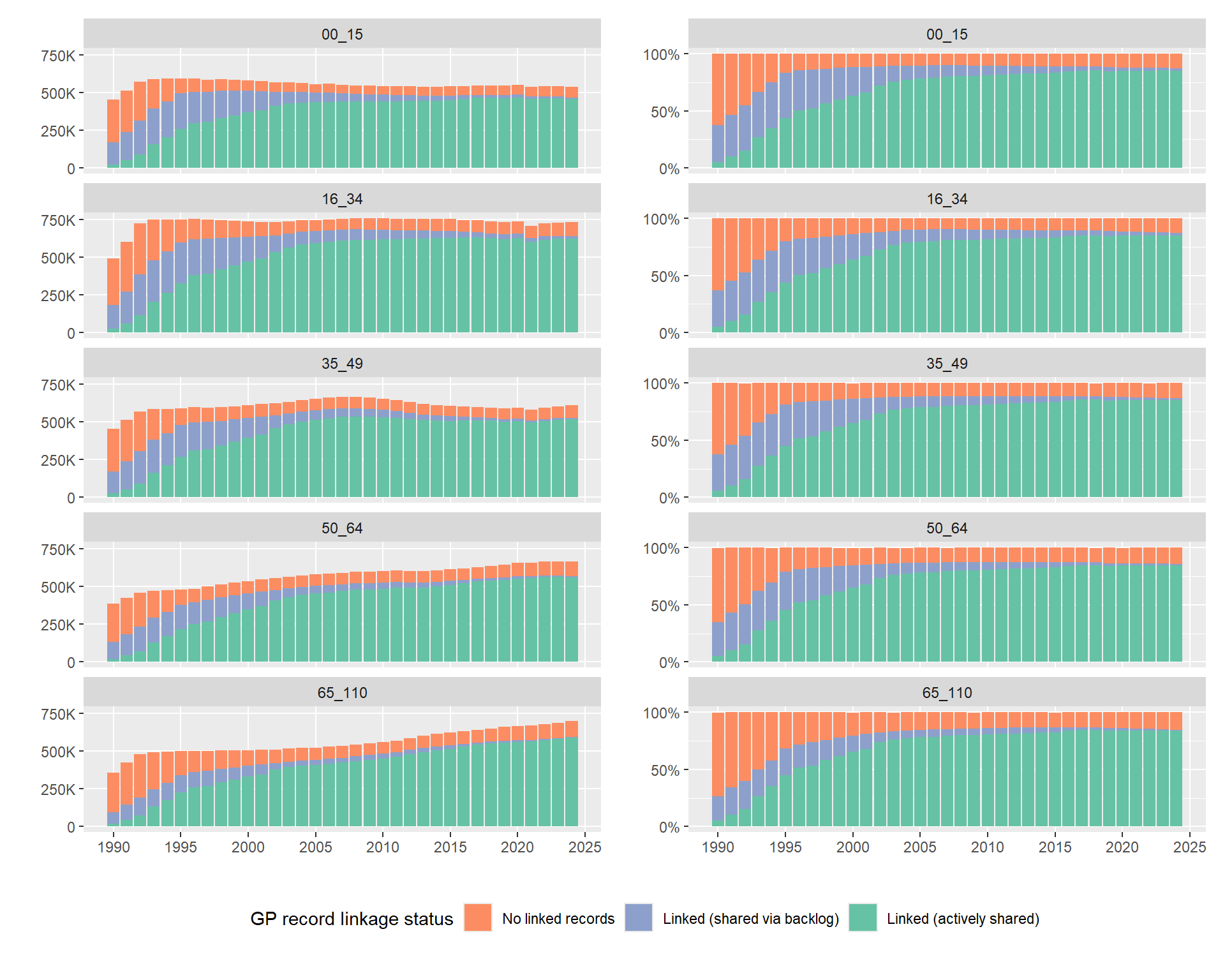

**Figure S3:** Mid-year population of Wales by SAIL-GP registration status from 1990 to 2024, stratified by Welsh Index of Multiple Deprivation (WIMD 2019).

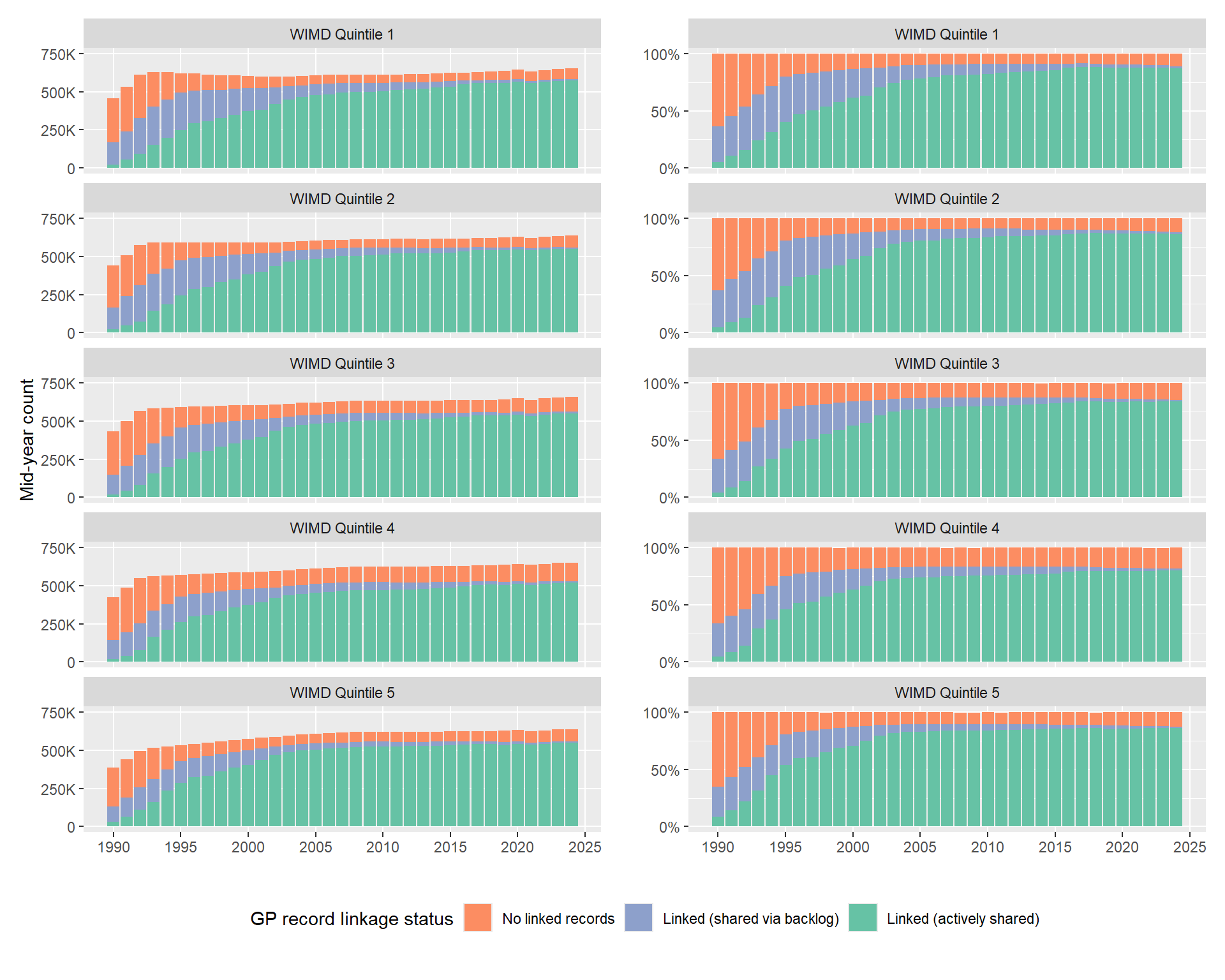

### S2. Trends of GP activity

For each GP activity category, we produced descriptive plots of the STL decomposition and autocorrelation. We have also included results from the longitudinal GAMM models: the fitted cyclic and thin-plate splines, which capture the seasonal variation and general longitudinal trend respectively, as well as the final fitted values from the models.

#### Consultation

**Figure S4:** STL decomposition of rates of averaged daily consultation between 2000 and 2024.

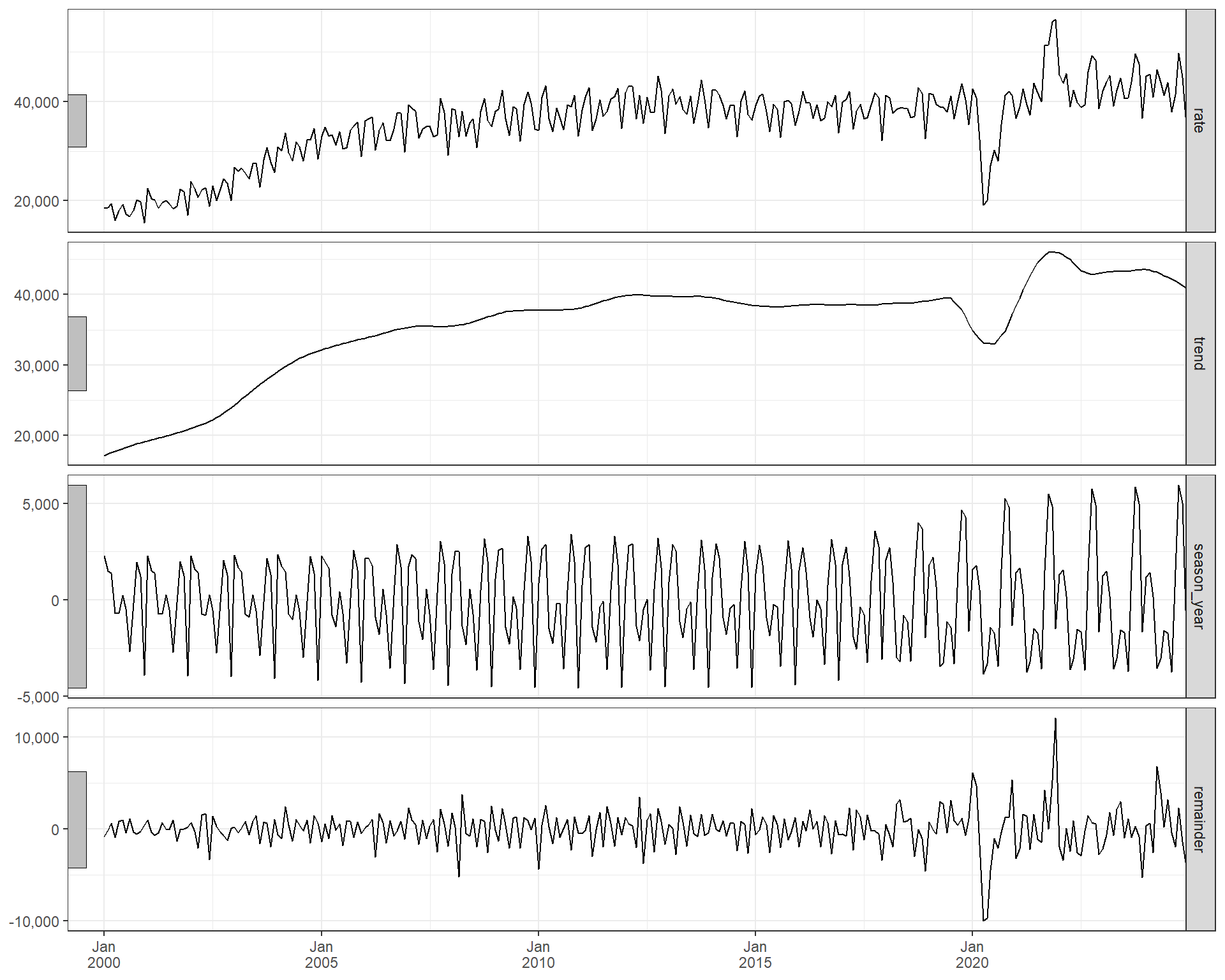

**Figure S5:** Autocorrelation Function (ACF) of consultation rates.

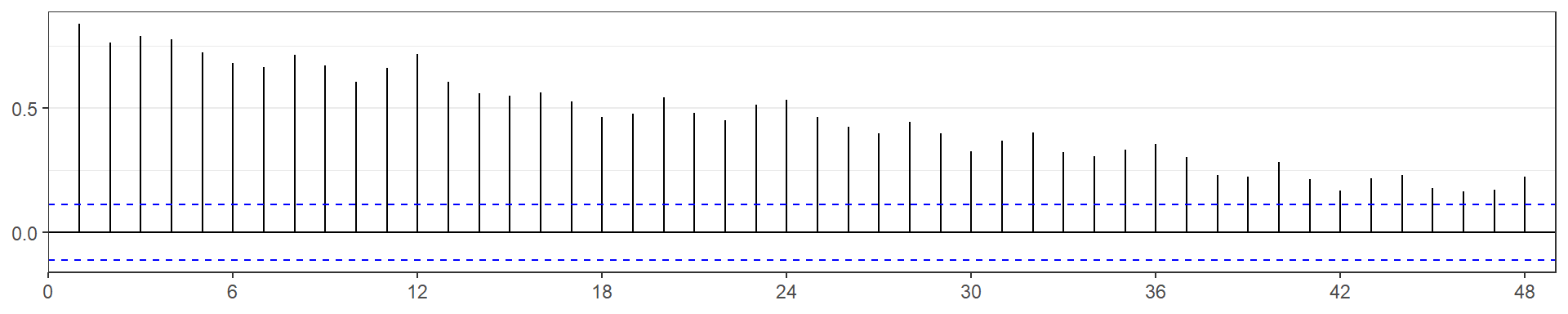

**Figure S6:** GAMM smoothing terms for calendar month and trend.

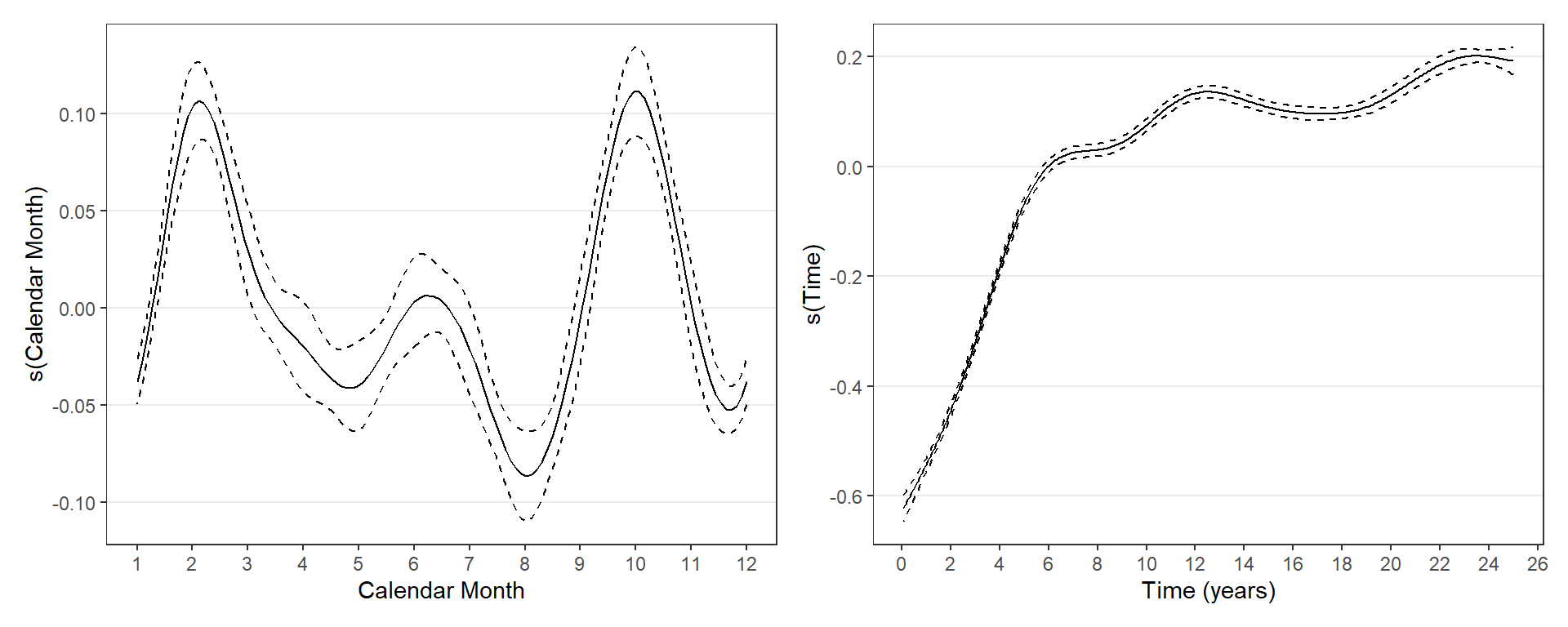

#### Prescription-only events

**Figure S7:** STL decomposition of rates of averaged daily prescription-only events between 2000 and 2024.

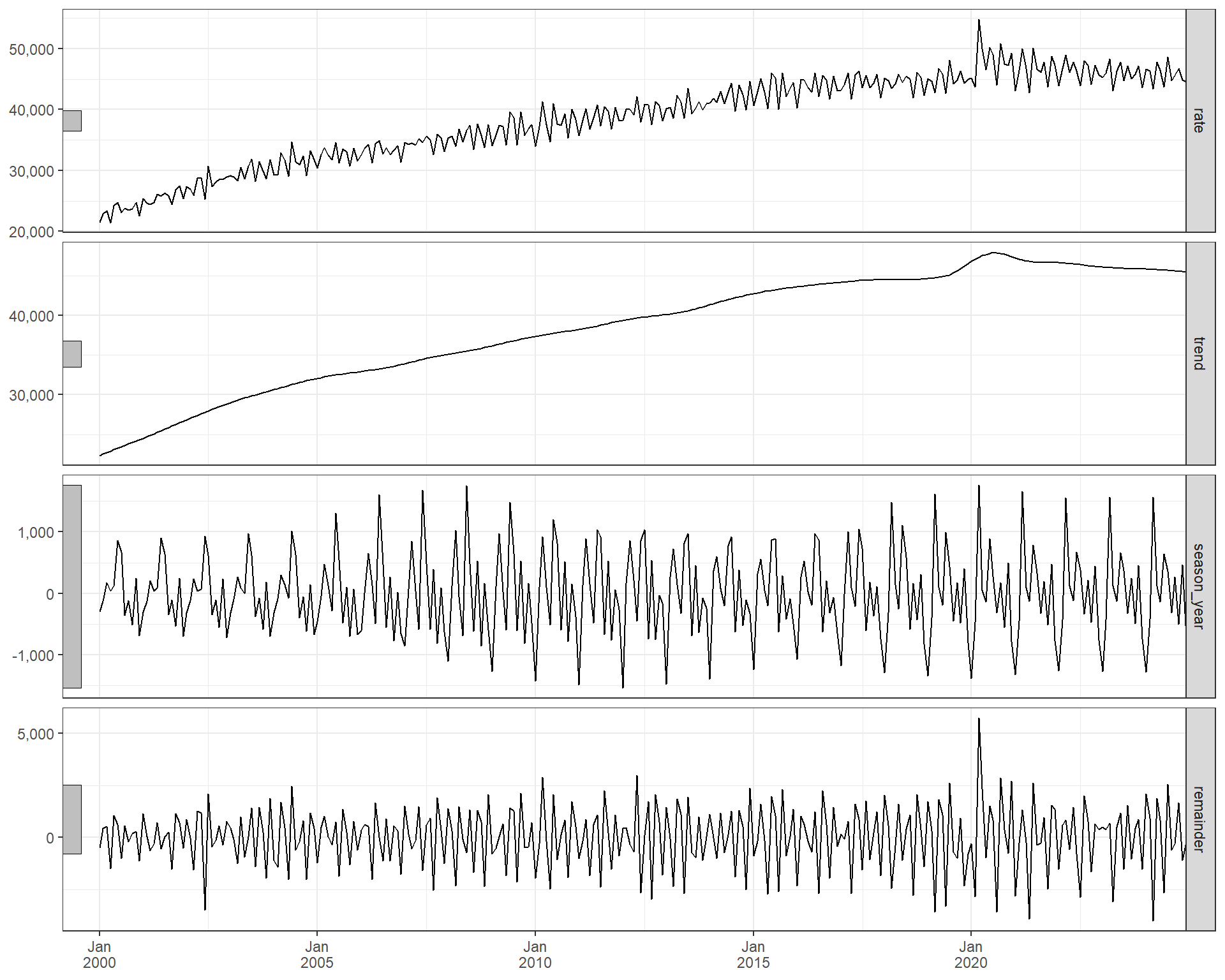

**Figure S8:** Autocorrelation Function (ACF) of prescription-only rates.

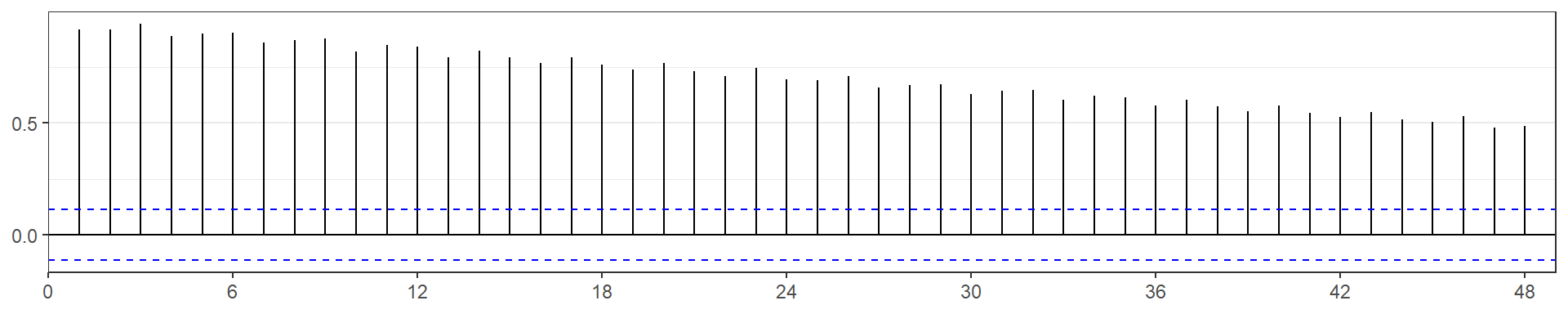

**Figure S9:** GAMM smoothing terms for calendar month and trend.

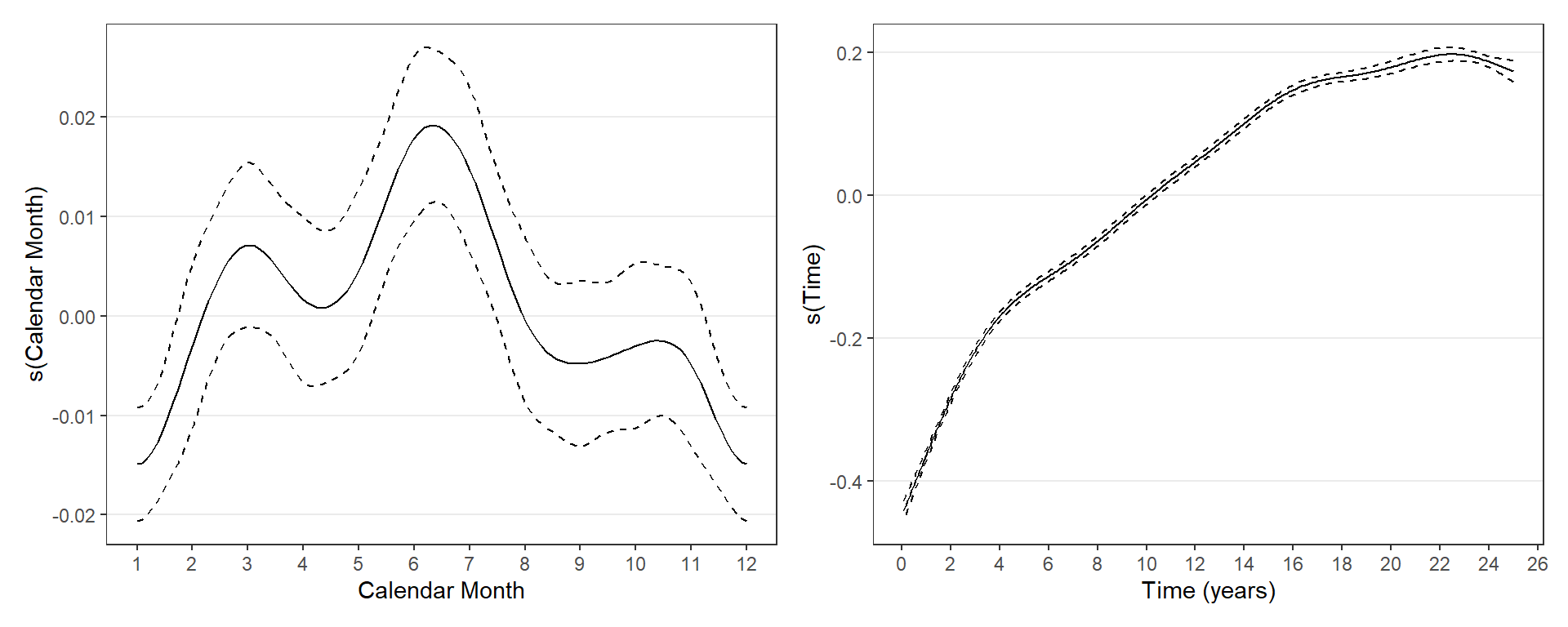

#### Vaccination

**Figure S10:** STL decomposition of rates of average daily rate of vaccinations per month between 2000 and 2024.

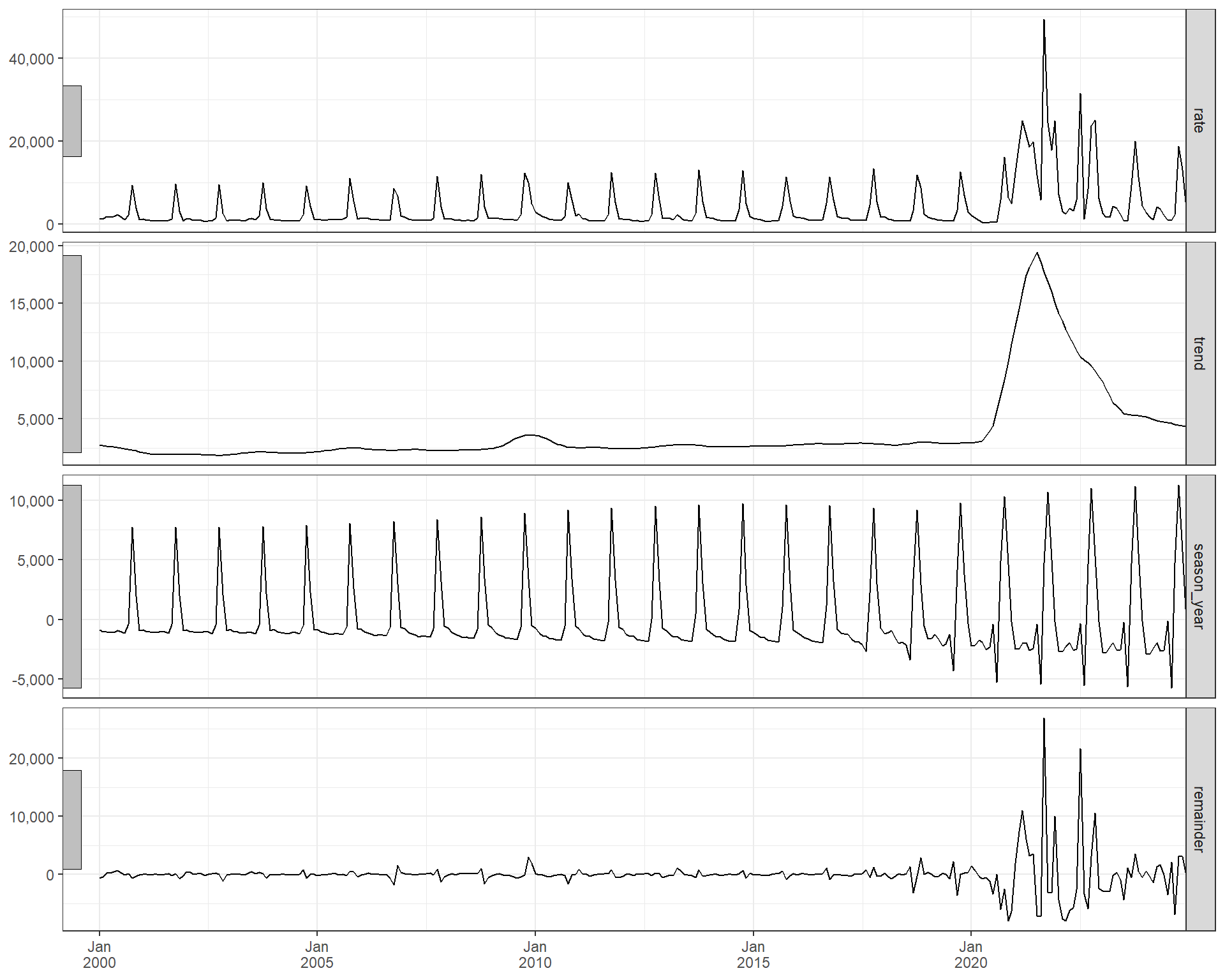

**Figure S11:** Autocorrelation Function (ACF) of vaccination rates.

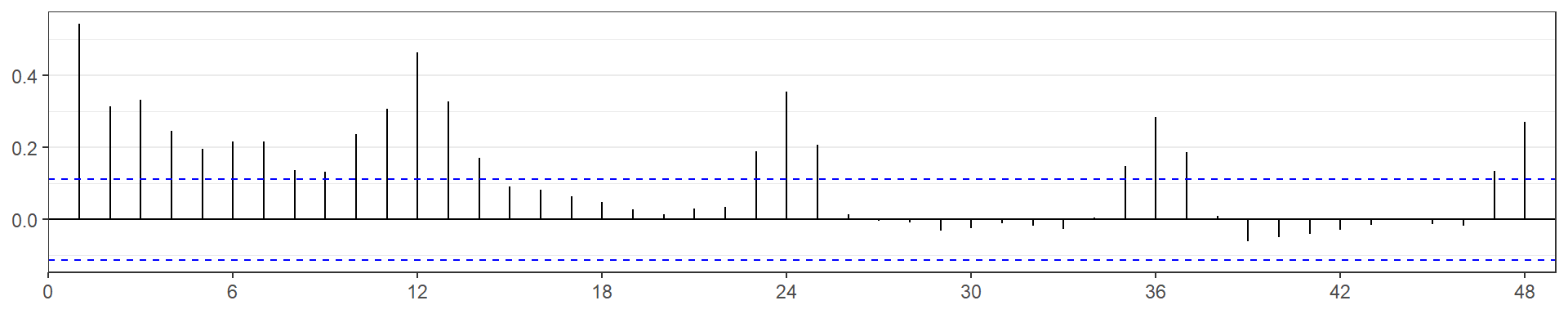

**Figure S12:** GAMM smoothing terms for calendar month and trend.

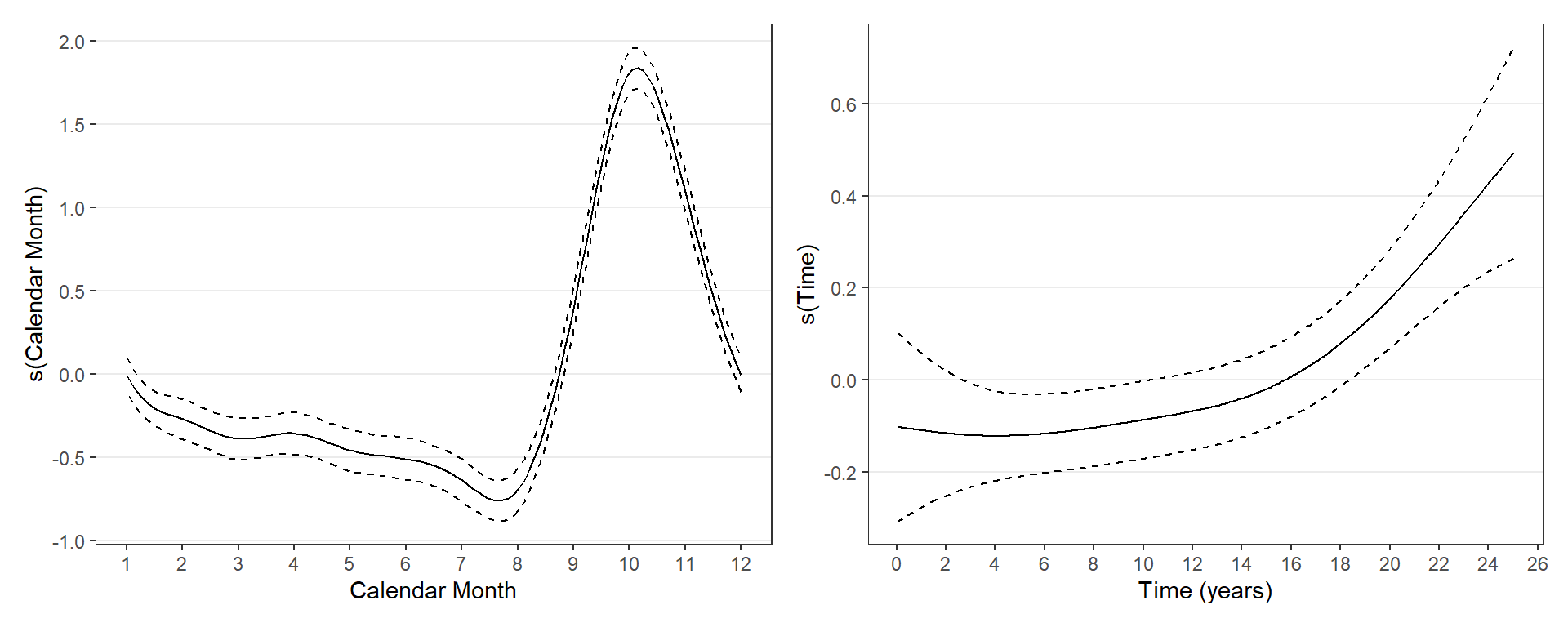

#### Patient review and monitoring

**Figure S13:** STL decomposition of rates average daily rate of patient review and patient monitoring events per month between 2000 and 2024.

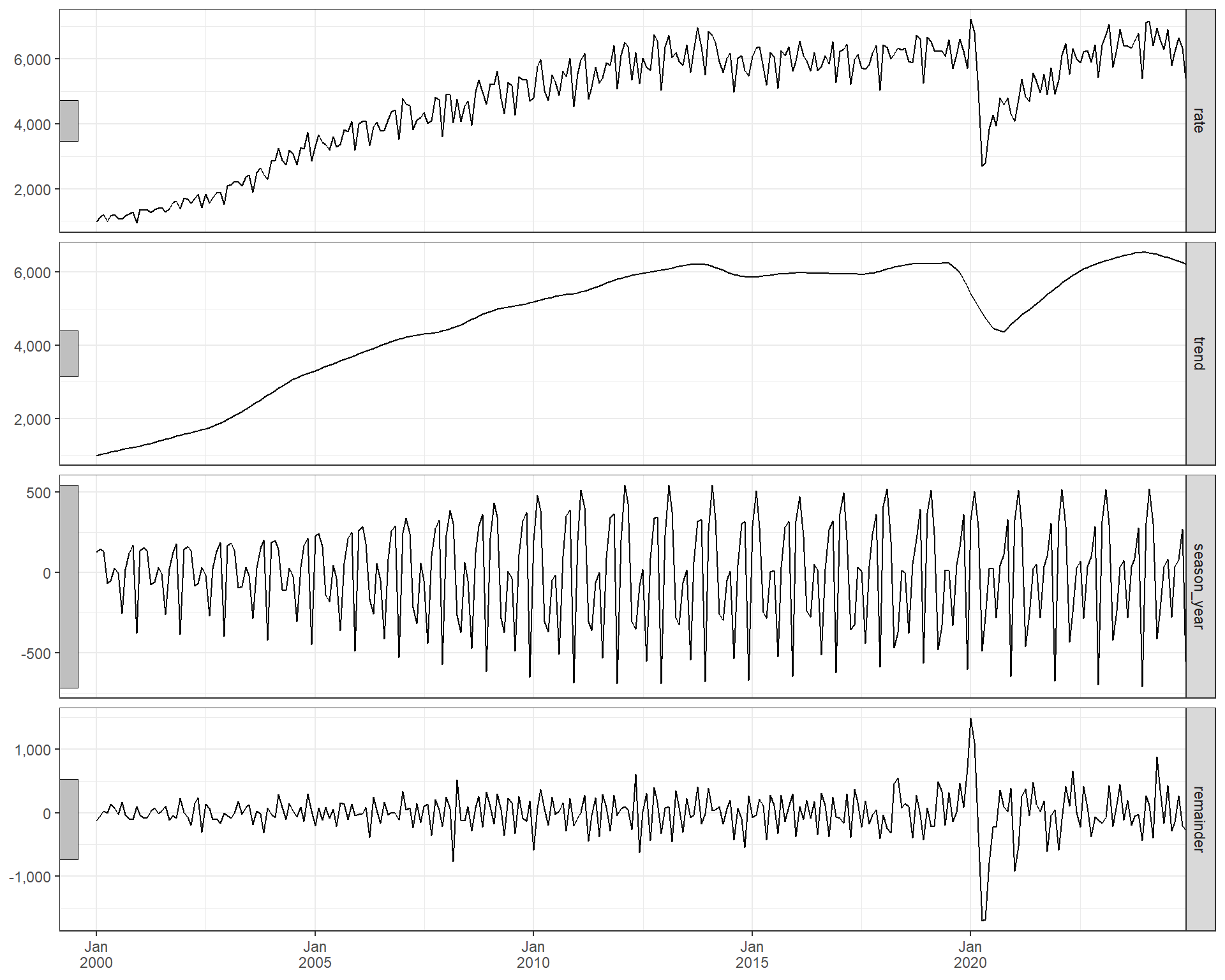

**Figure S14:** Autocorrelation Function (ACF) of patient review and patient monitoring rates.

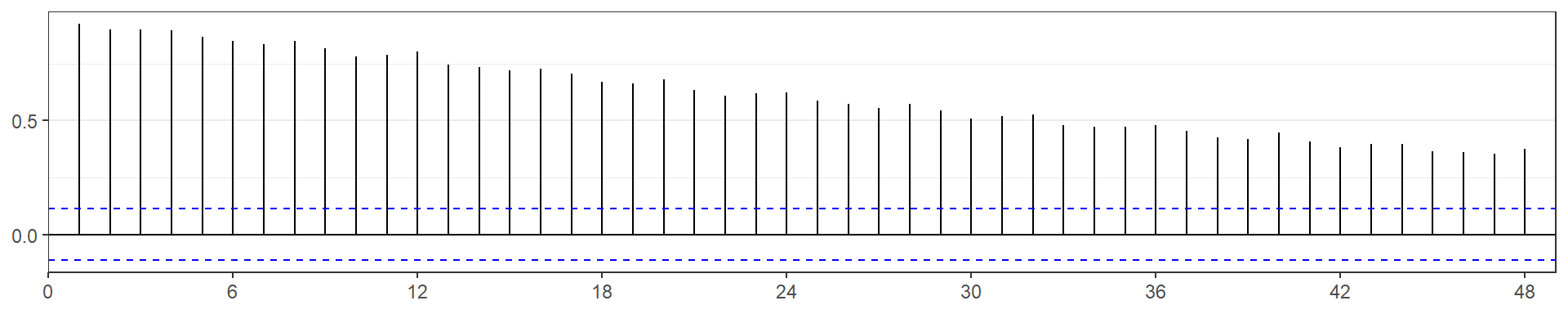

**Figure S15:** GAMM smoothing terms for calendar month and trend.

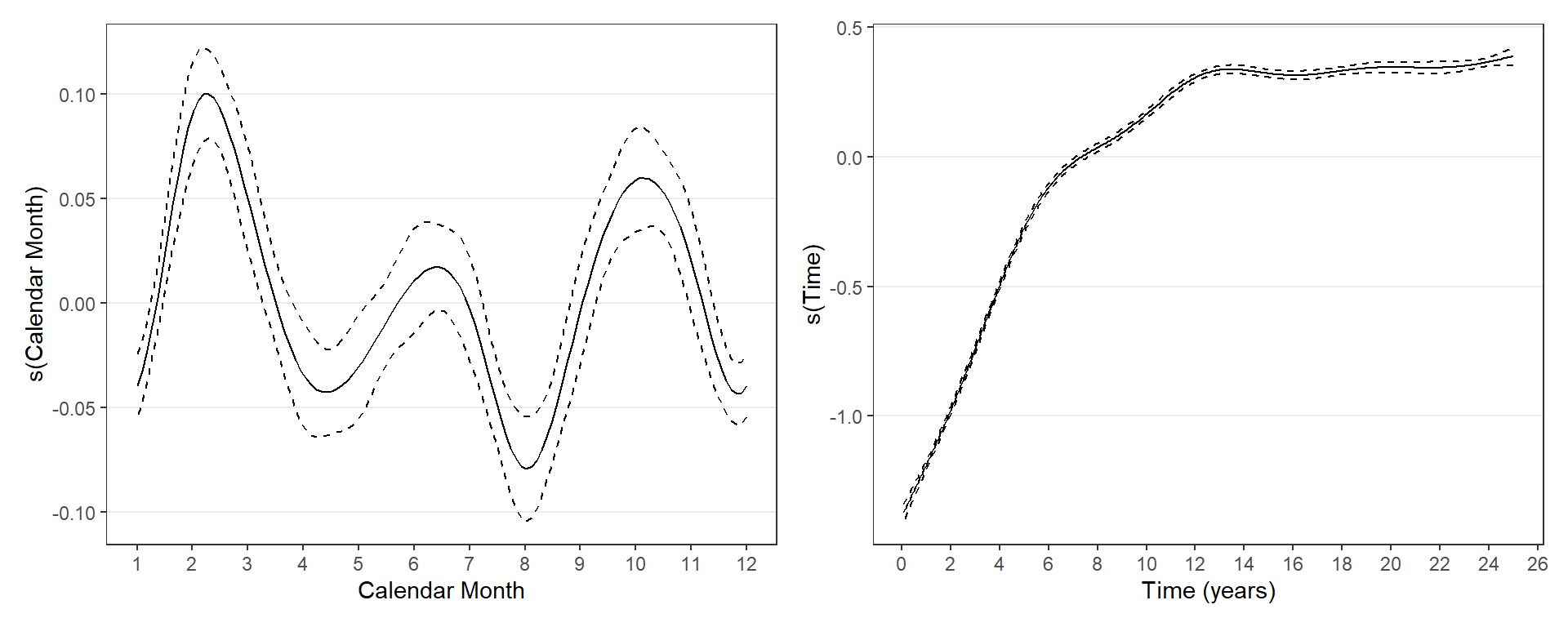

#### Screening and assessment

**Figure S16:** STL decomposition of rates of average daily screening and assessment events per month between 2000 and 2024.

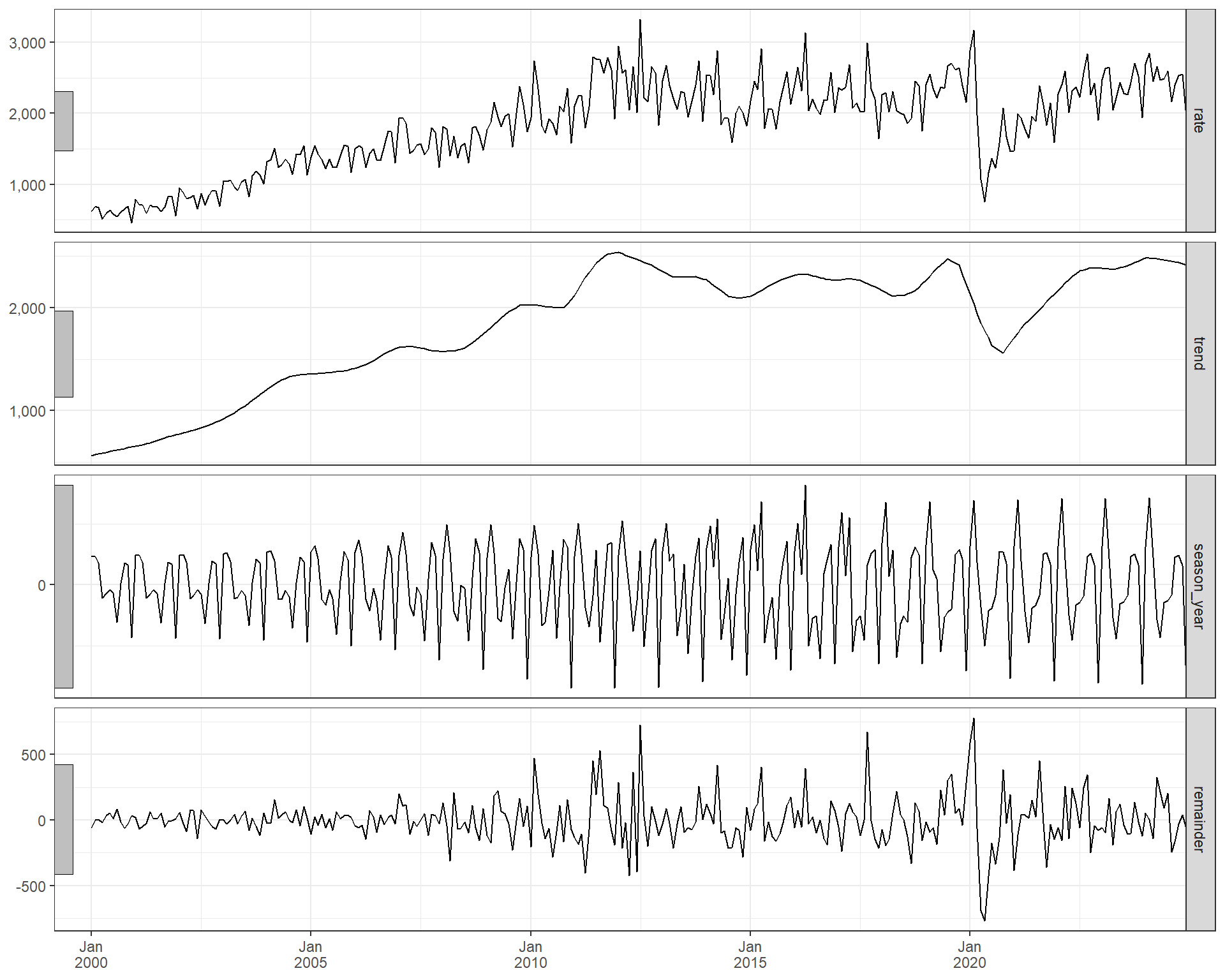

**Figure S17:** Autocorrelation Function (ACF) of screening and assessment rates.

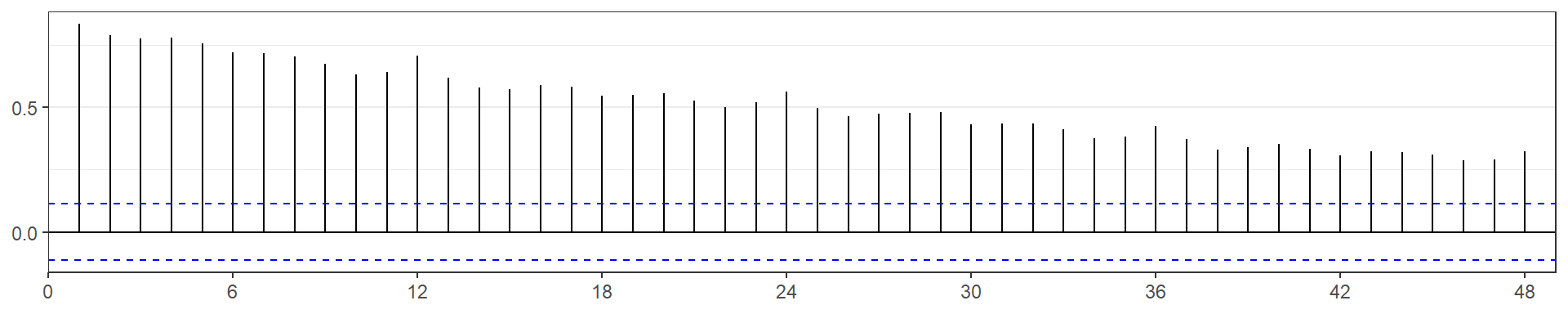

**Figure S18:** GAMM smoothing terms for calendar month and trend.

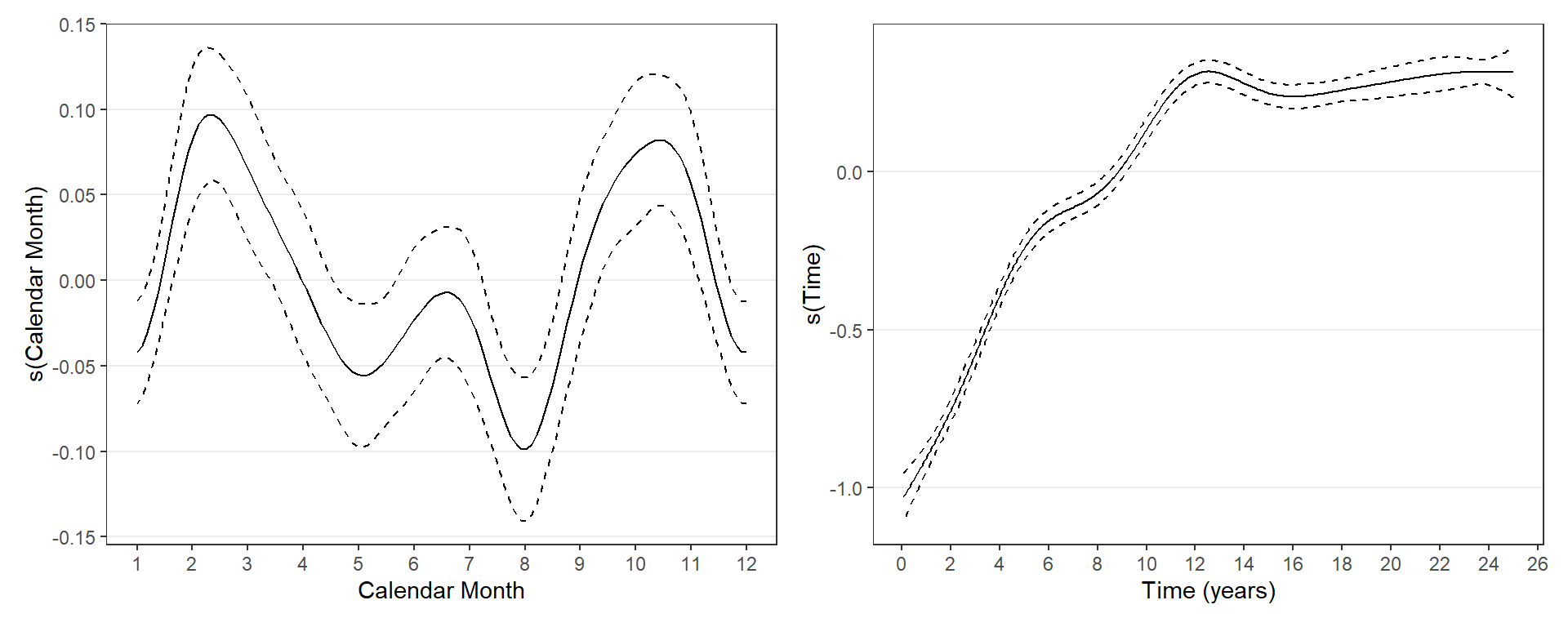

#### Summary of observed and expected trends across all GP activities

**Figure S19:** Observed and expected trends across all GP activities between 2000 and 2024.

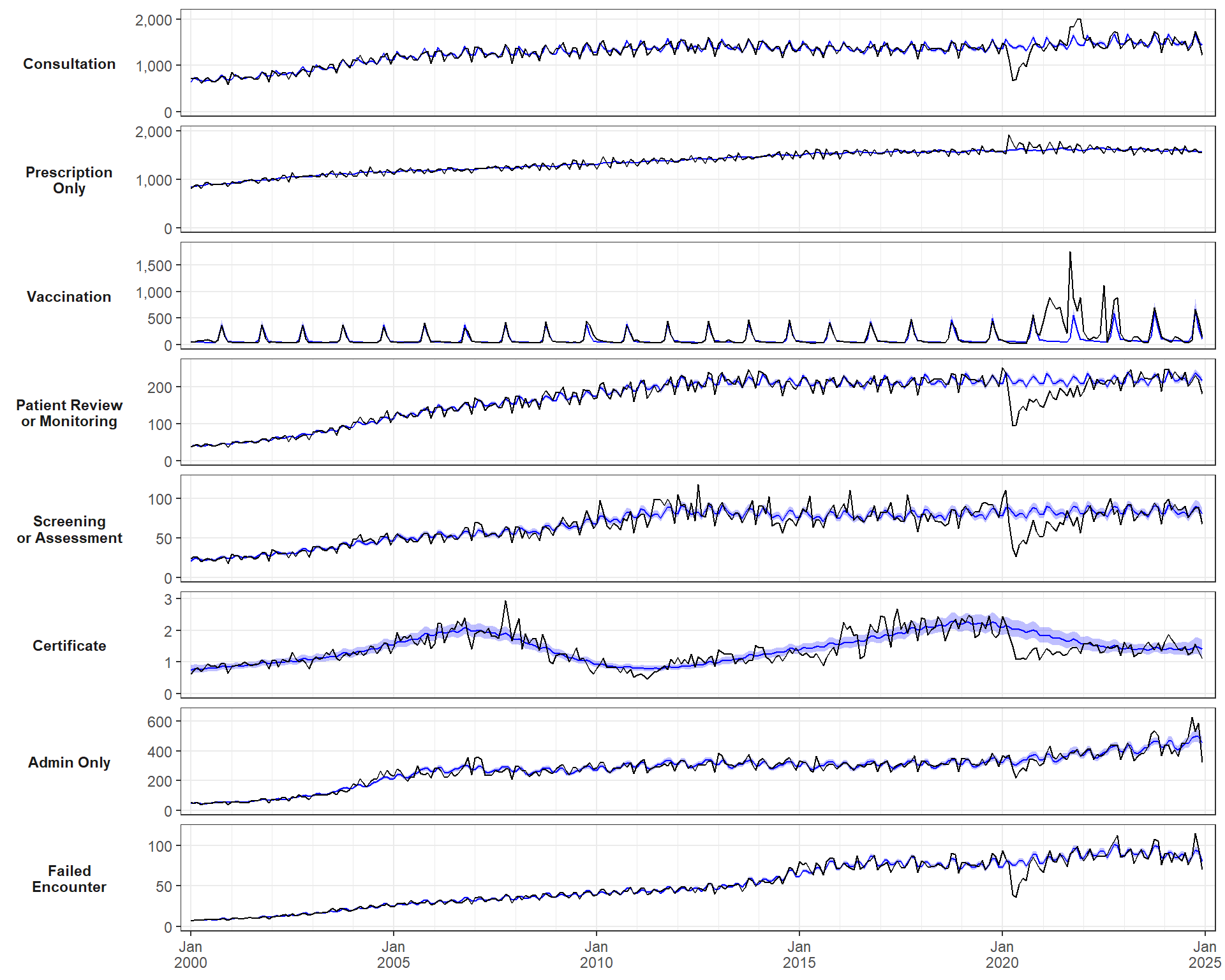
